# Predicting psychological symptoms when Facebook’s digital well-being features are used: A cross-sectional survey

**DOI:** 10.1101/2022.05.09.22274875

**Authors:** Tamara Barsova, Zi Gi Cheong, Ann R Mak, Jean CJ Liu

## Abstract

**Background:** Prior research has linked social media usage to poorer mental health. To address these concerns, social media platforms have introduced digital well-being tools to help users monitor their engagement. Nonetheless, little is known about the effectiveness of these tools.

**Objective:** In this study, we focused on Facebook to assess users’ awareness and usage of 6 Facebook well-being tools (‘Unfollow’, ‘Snooze’, ‘Off-Facebook Activity’, ‘Your Time on Facebook’, ‘Set Daily Reminders’, and ‘Notification Settings’). Additionally, we examined whether use of the tools was associated with better mental health outcomes.

**Methods:** We conducted a cross-sectional survey of 608 Facebook users. The survey comprised questions about: (i) baseline Facebook use; (ii) adoption of Facebook’s digital well-being tools; and (iii) participant demographics. These were used to predict the primary outcome measure – scores on the Depression, Anxiety and Stress Scale (DASS-21).

**Results:** Most participants (97%) knew about Facebook’s digital well-being tools, but each tool was used by only 17 to 55% of participants. In turn, use of two tools was associated with better well-being. Namely, although participants who spent more time on Facebook reported higher levels of depression, anxiety, and stress, those who managed their feed content or notifications (using ‘Unfollow’ or ‘Notification settings’) had lower scores on each of these measures. However, use of the ‘Snooze’, ‘Off-Facebook Activity’, ‘Your Time on Facebook’, or ‘Set Time Reminder’ features was not associated with lower depression, anxiety, or stress scores.

**Conclusions:** Of six of Facebook’s digital well-being tools, only two were associated with better user mental health. This underscores the complexity of designing social media platforms to promote user welfare. Consequently, we urge further research to understand the efficacy of various digital well-being tools.

**Trial Registration:** ClinicalTrials.gov NCT04967846

**Study registration:** NCT04967846: Social media effects on mental health (https://clinicaltrials.gov/ct2/show/study/NCT04967846)

## 1. Introduction

Over the past decade, social media platforms have been scrutinized for their potential impact on mental health. Amongst the general public, claims about social media harms have been widely publicized in both television documentaries [1] and whistle-blower accounts [2]. Within the academic literature, multiple studies have also linked social media usage to: symptoms of depression [3, 4], psychological distress [5], poorer well-being [5, 6], and lower self-esteem [7].

Two mechanisms have been proposed to explain why social media platforms may compromise mental health [1, 2]. First, the platforms allow users to compare themselves with celebrities or peers whose online posts portray more ideal lives than their own [8, 9]. This ‘upward social comparison’ may lead users to feel worse about themselves, placing them at risk for poorer mental health [8, 9]. Second, the platforms were designed to draw users’ attention for as long as possible [10, 11]. This may lead to excessive social media consumption, again impairing well-being [10, 11].

To address public concerns about these social media harms, the platforms’ app developers have introduced ‘digital well-being features’ to help users manage their engagement [12, 13]. Nonetheless, it remains unclear: (i) whether users know or use these features, and (ii) whether use of the features predicts better psychological well-being. Consequently, the present study examines these questions by focusing on Facebook as a case study.

### 1.1 Facebook’s digital well-being features

With 2.9 billion users worldwide, Facebook is the most widely-used social networking platform in the world [14]. Given its popularity, it has also been the focus of most research studies documenting the link between social media usage and poorer mental health [7, 9, 15]. As a result, Facebook developers consulted mental health experts and launched a series of digital well-being features, with the high-level goal that subsequent Facebook usage would be ‘intentional, positive and inspiring’ [16].

Facebook’s digital well-being features broadly address the two proposed mechanisms for social media harms. First, to minimize social comparison, several features allow users to curate the content that they see. For example, the ‘Unfollow’ option allows users to hide posts from selected friends, pages, or groups; while the ‘Snooze’ option hides these posts for a 30-day duration [17]. Finally, ‘Off-Facebook Activity’ allows users to customize how the platform integrates information from external apps to customize their feeds [17].

In the second category, a separate group of features enable users to monitor their usage patterns, curbing excessive use. For example, ‘Your Time on Facebook’ displays the amount of time a user has spent on Facebook over the past week, while ‘Set Daily Reminders’ notifies the user when a pre-determined cut-off has been reached (e.g., 45 mins of Facebook use) [17]. Finally, ‘Notification Settings’ allows users to manage the in-app notifications they receive, minimizing the amount of content that draws the user’s attention.

As no study has evaluated the efficacy of Facebook’s digital well-being tools, we conducted a cross-sectional survey to address two primary aims. First, we sought to document the extent to which Facebook users know and use the six outlined features: (i) Unfollow, (ii) Snooze, (iii) Off-Facebook Activity, (iv) Your Time on Facebook, (v) Set Daily Reminders, and (vi) Notification Settings. Second, we sought to replicate previous findings linking Facebook usage with poorer mental health, and to examine whether participants’ use of the well-being features was associated with better outcomes.

## 2. Materials and Methods

### 2.1 Study design and population

Participants were 608 Facebook users recruited from Amazon’s online panel (Mechanical Turk) in June 2021. All participants met the following eligibility criteria: (i) 21 years or older, (ii) proficient in English, (iii) based in the US, and (iv) with a positive track record on the platform (HIT approval rate > 95%, number of HITs approved > 500). Participants gave their written consent in accordance with the Declaration of Helsinki, and were given a nominal sum of USD $0.50 upon study completion. The study was approved by the Yale-NUS College Ethics Review Committee (2021-CERC-001) and was preregistered on ClinicalTrials.gov (NCT04967846).

### 2.2 Predictor variables

Predictor and outcome variables were measured through a 10-minute survey hosted on the website Qualtrics (https://osf.io/9z4dy/). Questions were written for a 7^th^ grade reading level, and were pilot-tested before the study.

#### 2.2.1 Baseline Facebook usage

The first set of questions captured participants’ baseline Facebook usage. Following studies linking Facebook use to mental health, participants estimated the number of hours they spent on Facebook over the past week [18].

To provide a context for these metrics, participants also reported how frequently they engaged in 9 Facebook activities (reading the news feed, posting status updates, posting photos, posting original content, browsing friends’ timelines, viewing friends’ photos, commenting on friends’ posts, sharing friends’ content, and using Facebook Messenger) [18]. These were rated using 7-point scales anchored with “never” and “more than once a day”.

#### 2.2.2 Awareness and adoption of Facebook well-being features

Central to this study, participants also reported their awareness and adoption of 6 Facebook digital well-being tools (Unfollow, Snooze, Off-Facebook Activity, Your Time on Facebook, Set Daily Reminders, and Notification Settings).

First, participants were shown screenshots of each feature and reported whether they had heard of it (Yes/No). If participants responded ‘yes’, they were then asked if they had used it (Yes/No). For features designed for repeated use (Unfollow, Snooze, Off-Facebook Activity, Your Time on Facebook), participants reported how frequently they used each feature (using a 5-point scale anchored with ‘never’ and ‘daily’).

#### 2.2.3 Demographics

As the final category of predictors, participants reported their: age, gender, race, religion, marital status, education, employment status, family income, household size, and living setting.

### 2.3 Outcome measures

#### 2.3.1 Mental health questionnaire

As an assay of mental health, participants completed the 21-item Depression, Anxiety, and Stress Scale (DASS-21) [19]. The DASS-21 is well-validated and widely used, consisting of 7 items per subscale: depression (e.g., “I couldn’t seem to experience any positive feelings at all”, “I found it difficult to work up the initiative to do things”; Cronbach’s alpha: 0.87), anxiety (e.g., “I was aware of dryness of my mouth”, “I felt I was close to panic” ; Cronbach’s alpha: 0.89), and stress (e.g., “I found it hard to wind down”, “I tended to over-react to situations”; Cronbach’s alpha: 0.89). Each item was rated with a 4-point scale (ranging from ‘0: did not apply to me at all’ to ‘3: applied to me very much or most of the time’), with scores summed and multiplied by two.

### 2.4 Statistical analysis

As part of data cleaning, we first verified that participants had read the questions through two verification items asking participants to check boxes as instructed (modelled after the widely-used CAPTCHA technique on the internet) [20]. Ten participants (1.6%) failed the verification and were removed from the dataset, resulting in a final sample of 598 participants. We then summarized participants’ baseline characteristics using counts (with percentages) and medians (with interquartile ranges).

As the primary analyses, we ran a series of linear regression models, using each DASS-21 subscale score (depression, anxiety, and stress) as an outcome measure. In the first model, we sought to replicate the oft-reported link between one’s duration of Facebook use and poorer mental health [8]. To this end, we entered the number of hours participants spent using Facebook as a predictor, log-transformed to achieve linearity (Model 1).

In the second model, we addressed the study’s primary aim, examining whether adoption of Facebook’s well-being features predicted better mental health (having controlled for duration of Facebook use). Correspondingly, Model 1 was repeated with 6 additional predictors that coded for the use of each feature (Unfollow, Snooze, Off-Facebook Activity, Your Time on Facebook, Set Daily Reminders, Notification Settings; Model 2). For each predictor, non-usage was coded as ‘0’ and usage as ‘1’.

Finally, we assessed the robustness of our findings by repeating Model 2 with the inclusion of demographic variables (age, gender, race, religion, marital status, education, employment, family income, household size, living setting) (Model 3).

Across the models, the type 1 decision-wise error rate was controlled at α = 0.05, with adequate statistical power (0.80) to detect small effect sizes (f^2^ = 0.05). All statistical analyses were carried out on SPSS 27 (IBM Corp, Armonk, NY) and R4.0.3 (R Core Team, Vienna, Austria).

## 3. Results

### 3.1 Participant characteristics and baseline Facebook usage

Participant demographics were comparable to that of US Facebook users [21]: namely, 3 in 4 participants (75.7%) were aged 45 years or below, and slightly over half the participants (60.2%) self-identified as male (Table 1).

**Table 1.** Baseline characteristics of survey respondents.

| Characteristic | N | (%) |
| --- | --- | --- |
| *Age group* |  |  |
| · <35 years | 309 | (51.6) |
| 21-25 | 45 | (7.5) |
| 26-35 | 264 | (44.1) |
| · ≥35 years | 289 | (48.4) |
| 36-45 | 144 | (24.1) |
| 46-55 | 83 | (13.9) |
| 56 and above | 62 | (10.4) |
| *Gender* |  |  |
| · Female | 237 | (39.6) |
| · Male | 360 | (60.2) |
| · Non-binary / third gender | 1 | (0.2) |
| *Race* |  |  |
| · White | 475 | (79.4) |
| · Black or African American | 84 | (14.0) |
| · Other | 39 | (6.5) |
| Asian | 20 | (3.3) |
| American Indian or Alaska Native | 9 | (1.5) |
| Two or more races | 6 | (1.0) |
| Other | 4 | (0.7) |
| *Religion* |  |  |
| · No religion | 81 | (13.5) |
| · Christianity (Protestant) | 370 | (61.9) |
| · Christianity (Catholic) | 107 | (17.9) |
| · Others | 121 | (20.2) |
| Buddhism | 17 | (2.8) |
| Hinduism | 10 | (1.7) |
| Islam | 3 | (0.5) |
| Others | 10 | (1.7) |
| *Marital status* |  |  |
| · Married / partnered | 464 | (77.6) |
| · Single | 116 | (19.4) |
| · Others | 18 | (3.0) |
| Divorce | 16 | (2.7) |
| Separated | 2 | (0.3) |
| *Education level* |  |  |
| · Less than high school | 1 | (0.2) |
| · High school diploma or equivalent | 26 | (4.3) |
| · Associate degree | 26 | (4.3) |
| · Bachelor's degree | 398 | (66.6) |
| · Some college but no degree | 37 | (6.2) |
| · Postgraduate degree (Master's, Doctoral) | 68 | (11.4) |
| · Professional degree (JD, MD) | 42 | (7.0) |
| *Employment status* |  |  |
| · Full time: 40 hours or more per week | 507 | (84.8) |
| · Not full time | 91 | (15.1) |
| Part time: up to 39 hours per week | 42 | (7.0) |
| Self-employed | 25 | (4.2) |
| Retired | 8 | (1.3) |
| Unemployed looking for work | 7 | (1.2) |
| Unemployed not looking for work | 5 | (0.8) |
| Unable to work | 2 | (0.3) |
| Student | 2 | (0.3) |
| *Family income level* |  |  |
| · Below \$30,000 | 68 | (11.4) |
| · \$30,000 - \$49,999 | 149 | (24.9) |
| · \$50,000 - \$74,999 | 212 | (35.5) |
| · \$75,000 - \$99,999 | 115 | (19.2) |
| · \$100,000 or above | 54 | (9.0) |
| *Household size* |  |  |
| · 1 | 57 | (9.5) |
| · 2 | 88 | (14.7) |
| · 3 | 219 | (36.6) |
| · 4 | 185 | (30.9) |
| · 5 or more | 49 | (8.2) |
| *Living setting* |  |  |
| · Large city | 195 | (32.6) |
| · Suburb | 117 | (19.6) |
| · Rural | 107 | (17.9) |
| · Large town | 94 | (15.7) |
| · Small town | 84 | (14.0) |
| · Others | 1 | (0.2) |
| *Average frequency of Facebook use* |  |  |
| · Never | 15 | (2.5) |
| · Once a week | 21 | (3.5) |
| · 2-3 times a week | 52 | (8.7) |
| · 4-6 times a week | 89 | (14.9) |
| · Once a day | 130 | (21.8) |
| · Multiple times a day | 289 | (48.5) |

In terms of baseline Facebook usage, participants reported using the platform for a median of 3 hours in the preceding week (IQR = 6 hours). Nearly 1 in 2 participants (48.3%, 95% CI: 44.3%-52.3%) accessed Facebook multiple times a day, while another 1 in 5 (21.7%, 95% CI: 18.4%-25.0%) logged in at least once daily (Table 1). On Facebook, participants were most likely to view a friend’s photo(s) or to read the news feed (with 7 in 10 participants engaging in each of these at least once a week; Figure 1).

**Figure 1.**
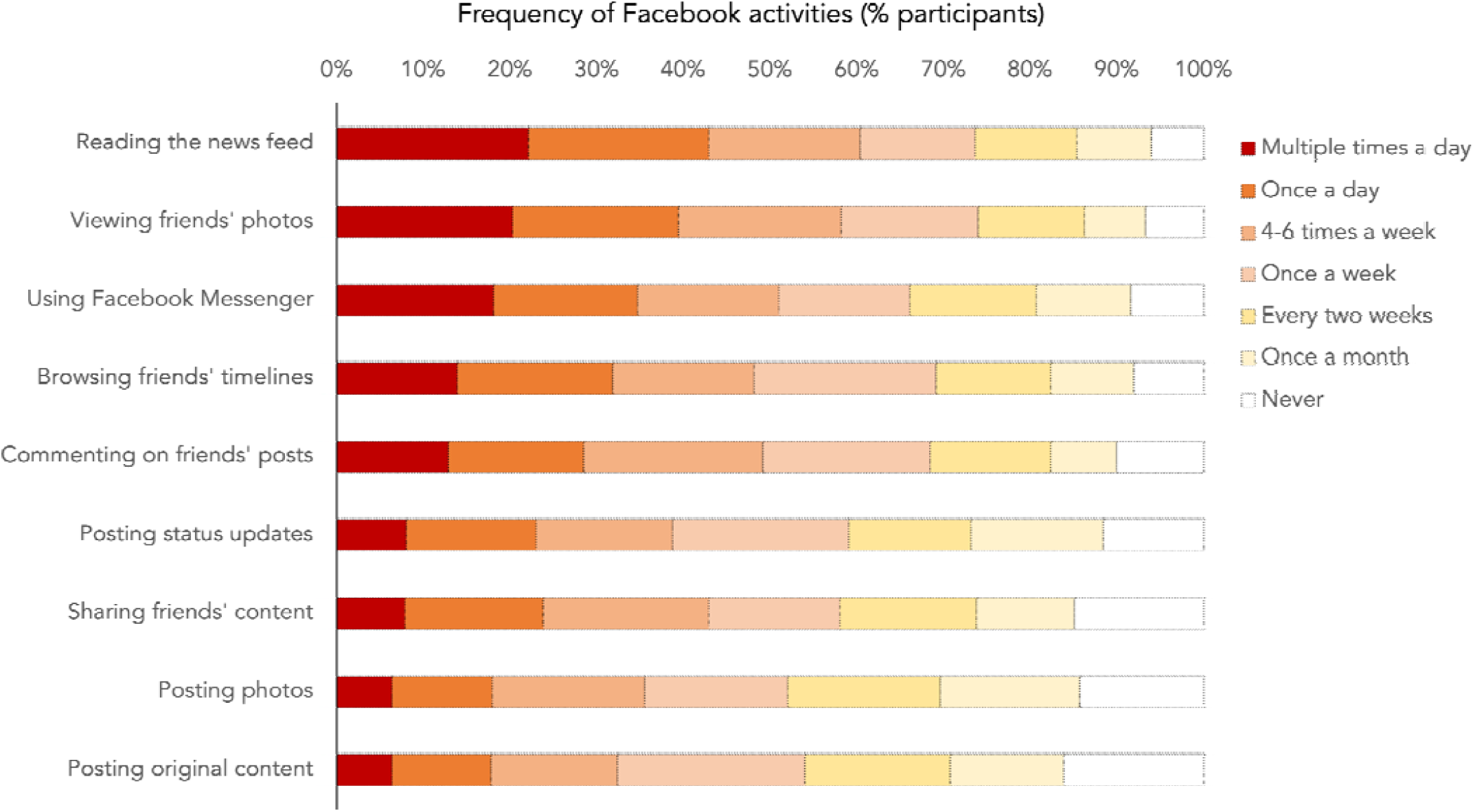
For each of 9 Facebook activities, participants rated whether they used it: (i) multiple times a day, (ii) once a day, (iii) 4-6 times a week, (iv) once a week, (v) every two weeks, (vi) once a month, or (vii) never. Each horizontal bar indicates the percentage of participants who chose each option.

### 3.2 Awareness and use of Facebook well-being features

Overall, most participants 97% (95% CI: 95.6% – 98.4%) were aware of at least one of Facebook’s well-being features (Figure 2). However, awareness levels differed across features: for example, while 85% (95% CI: 82.1% – 87.9%) had heard of Notification Settings, only 43.3% (95% CI: 39.7% – 47.3%) of participants knew about Your Time on Facebook.

**Figure 2.**
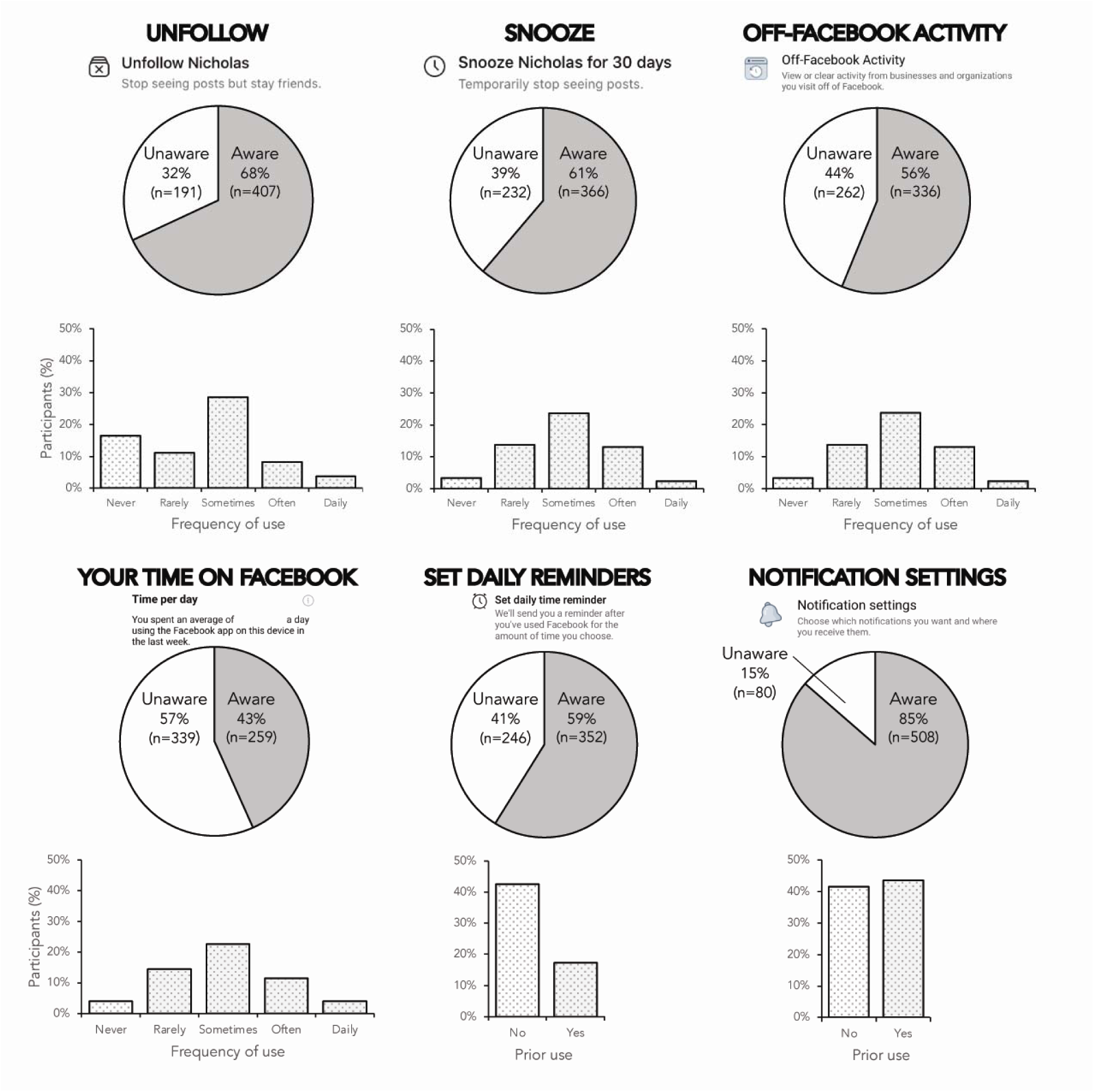
Participants indicated their awareness and usage of Facebook’s in-app digital well-being tools (Unfollow, Snooze, Off-Facebook Activity, Your Time on Facebook, Set Daily Reminders, Notification Settings).

In terms of usage, 1 in 2 participants had used: Snooze (55.5%, 95% CI: 51.5% – 59.5%), Off-Facebook Activity Tracker (52.7%; 95% CI: 48.7% – 56.7%), Your Time on Facebook (52.6%, 95% CI: 48.6% – 56.6%), and Unfollow (51.7%, 95% CI: 47.7% – 55.7%). Less than half had adjusted Notification Settings (43.5%, 95% CI: 39.5% – 47.5%), and fewer still had used the Set Time Reminder (17.4%, 95% CI: 14.4% – 20.4%). Where repeated use of the features was possible, participants were most likely to report using them ‘sometimes’, on an ad-hoc rather than routine basis (based on the median ratings for: Snooze, Off-Facebook Activity, Your Time on Facebook, and Unfollow) (Figure 2).

### 3.3 Use of Facebook well-being features and psychological symptoms

For the primary research question, we sought to predict participants’ depression, anxiety, and stress scores as a function of whether they used Facebook’s well-being features.

#### 3.3.1 Depression

In terms of depression, we first replicated the well-documented association between Facebook usage and depression symptoms: namely, the more time participants spent using Facebook, the higher their depression scores (Model 1: β=2.754, *P*<0.001; Model 2: β=1.357, *P*<0.001; Model 3: β=1.586, *P*<0.001) (Table 2).

Factoring whether participants used Facebook’s well-being features significantly increased the amount of variance in depression scores accounted for – from 8.7% (Model 1) to 28.2% (Model 2; F(6, 587)=28.015, R^2^=0.203, *P*<0.001). While use of the Notification Settings feature (β= -1.579, *P*<0.003) and the Unfollow button (β= -1.319, *P*=0.22) was associated with lower depression scores, use of Off-Facebook Activity (β=4.905, *P*<0.001) and the Snooze function (β=2.337, *P*<0.001) was associated with higher depression scores.

There was no significant association between depression scores and participants’ use of either Your Time on Facebook or Set Time Reminder (smallest *P*=0.48).

Each of these findings was robust, and persisted even when demographic variables were controlled for (in Model 3).

#### 3.3.2 Anxiety

As with depression symptoms, duration of Facebook use predicted increased anxiety scores (Model 1: β=4.331, *P*<0.001; Model 2: β=2.270, *P*<0.001; Model 3: β=2.1846, *P*<0.001) (Table 3).

**Table 3.**
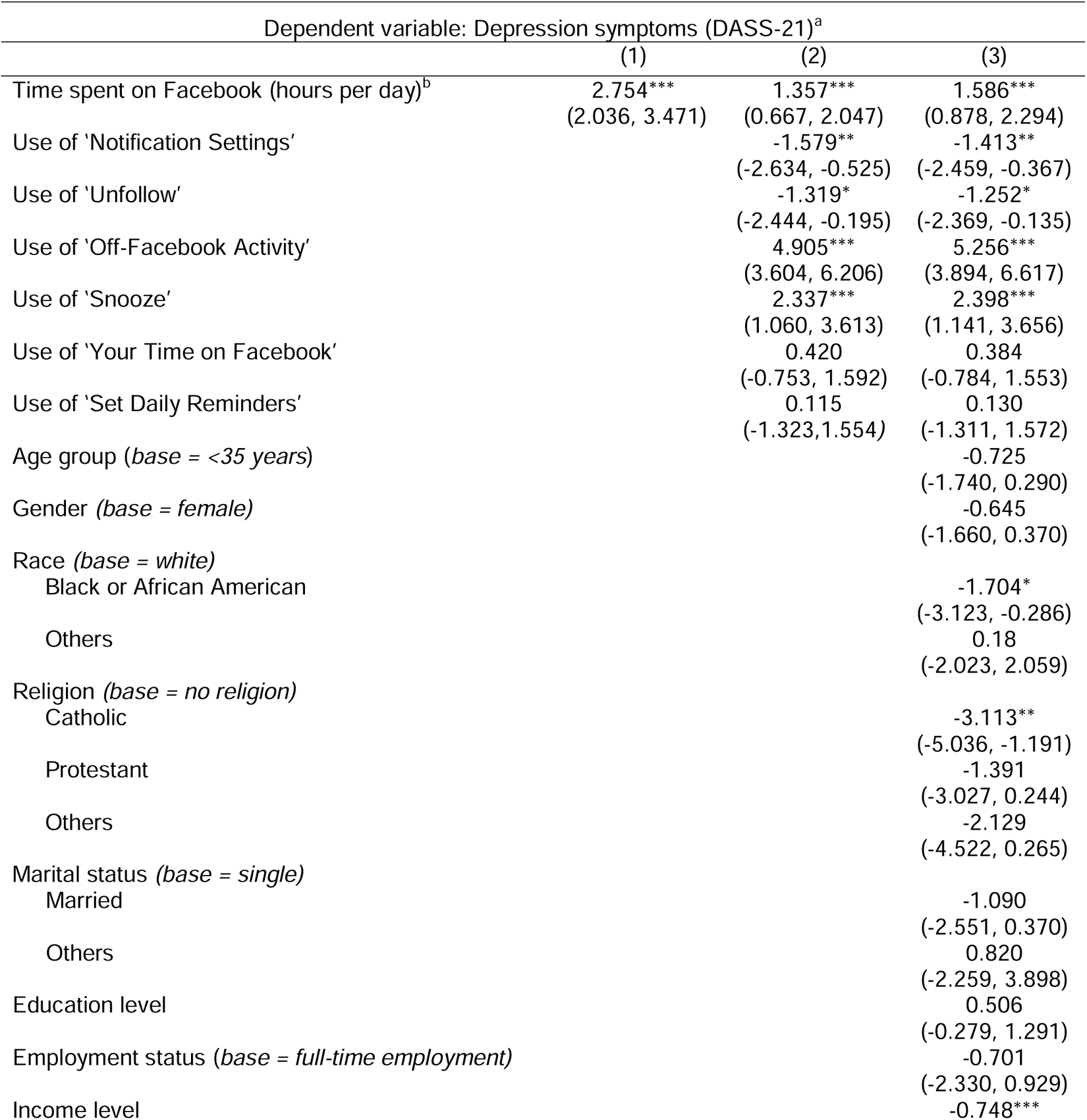
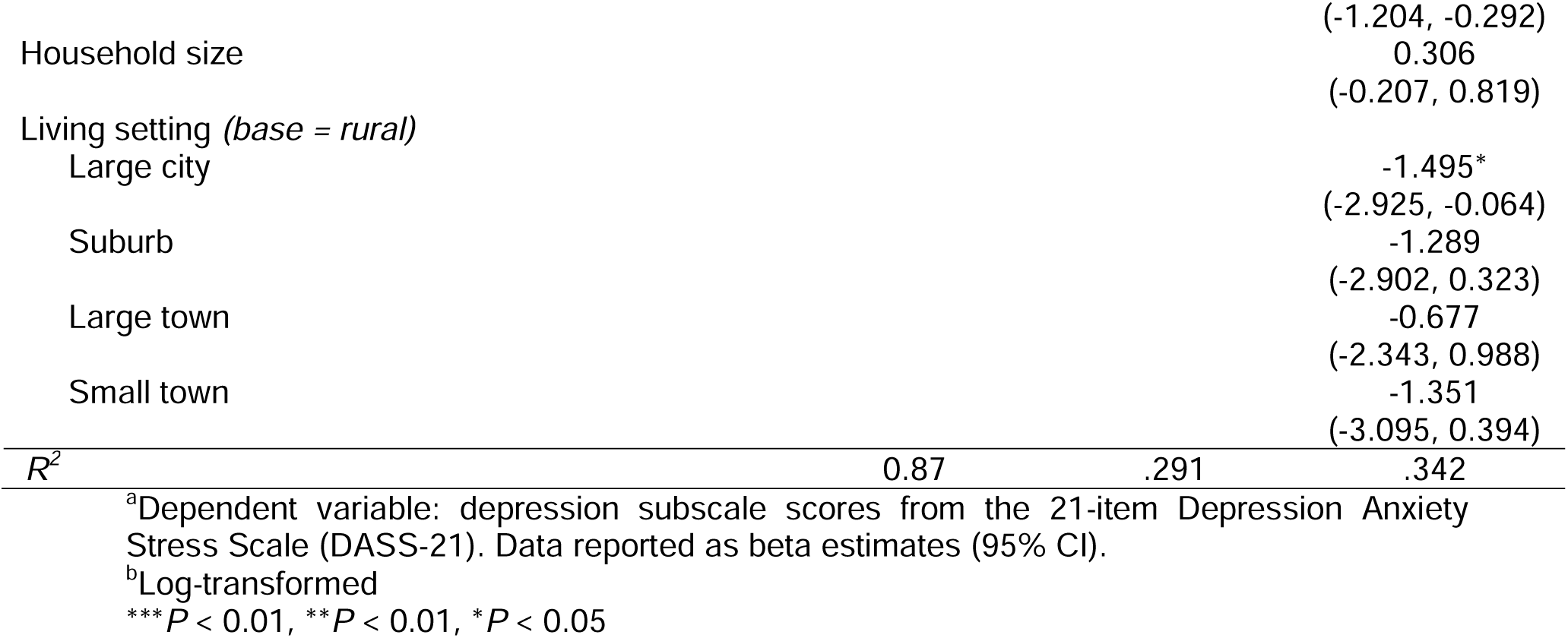
Predicting depression symptoms as a function of Facebook usage patterns.

Again, the inclusion of variables that coded for participants’ use of Facebook’s well-being features increased the amount of variance accounted for (from 13.8% in Model 1 to 40.3% in Model 2; F(6, 587)=43.362, R^2^=0.265, *P*<0.001). Namely, while use of the Notification Shortcut Bar (β= -2.387, *P*<0.001) and Unfollow functions (β= -1.603, *P*=0.015) emerged as protective factors, use of Off-Facebook Activity (β=6.760, *P*<0.001) and the Snooze functions (β=3.134, p<0.001) predicted higher anxiety. We found no evidence that anxiety scores were linked to use of either Your Time on Facebook or Set Time Reminder (smallest *P*=0.07). Each of these findings persisted when we controlled for demographic variables in Model 3.

#### 3.3.3 Stress

Finally, time spent on Facebook was again linked to increased stress scores (Model 1: β=3.851, *P*<0.001; Model 2: β=1.825, *P*<0.00; Model 3: β=2.103, *P*<0.001) (Table 4).

**Table 4.** Predicting anxiety symptoms as a function of Facebook usage patterns.

| Dependent variable: Anxiety symptoms (DASS-21)<sup>a</sup> |  |  |  |
| --- | --- | --- | --- |
|  | (1) | (2) | (3) |
| Time spent on Facebook (hours per day)<sup>b</sup> | 4.331***<br>(3.459, 5.203) | 2.270***<br>(1.479, 3.062) | 2.184***<br>(1.376, 2.992) |
| Use of 'Notification Settings' |  | -2.387***<br>(-3.596, -1.177) | -2.064***<br>(-3.257, -0.871) |
| Use of 'Unfollow' |  | -1.603*<br>(-2.893, -0.314) | -1.593*<br>(-2.868, -0.319) |
| Use of 'Off-Facebook Activity' |  | 6.760***<br>(5.268, 8.252) | 6.151***<br>(4.598, 7.705) |
| Use of 'Snooze' |  | 3.134***<br>(1.670, 4.598) | 3.326***<br>(1.891, 4.760) |
| Use of 'Your Time on Facebook' |  | 1.225<br>(-0.120, 2.569) | 0.884<br>(-0.449, 2.217) |
| Use of 'Set Daily Reminders' |  | 0.327<br>(-1.323, 1.977) | 0.078<br>(-1.556, 1.723) |
| Age group (*base* = <35 years) |  |  | -0.996<br>(-2.155, 0.162) |
| Gender (*base* = *female*) |  |  | -1.165*<br>(-2.323, -0.006) |
| Race (*base* = *white*) |  |  |  |
| Black or African American |  |  | -1.227<br>(-2.846, 0.391) |
| Others |  |  | -0.503<br>(-2.832, 1.826) |
| Religion (*base* = *no religion*) |  |  |  |
| Catholic |  |  | -1.864<br>(-4.057, 0.329) |
| Protestant |  |  | 0.027<br>(-1.839, 1.894) |
| Others |  |  | -0.968<br>(-3.699, 1.763) |
| Marital status (*base* = *single*) |  |  |  |
| Others |  |  | -3.877*<br>(-7.390, -0.365) |
| Married |  |  | -0.742<br>(-2.409, 0.924) |
| Education level |  |  | 0.879<br>(-0.016, 1.774) |
| Employment status (*base* = *full-time employment*) |  |  | -1.606<br>(-3.465, 0.254) |
| Income level |  |  | -0.737**<br>(-1.258, -0.217) |
| Household size |  |  | 0.411,<br>(-0.175, 0.996) |
| Living setting (*base* = *rural*) |  |  |  |
| Large city |  |  | -1.958*<br>(-3.590, -0.326) |
| Suburb |  |  | -2.398*<br>(-4.238, -0.558) |
| Large town |  |  | -0.997<br>(-2.897, 0.903) |
| Small town |  |  | -1.385<br>(-3.375, 0.605) |
| $R^2$ | 0.138 | 0.403 | 0.452 |

<sup>a</sup>Dependent variable: Anxiety subscale scores from the 21-item Depression Anxiety Stress Scale (DASS-21). Data reported as beta estimates (95% CI).
<sup>b</sup>Log-transformed
\*\*\*P < 0.01, \*\*P < 0.01, \*P < 0.05

**Table 5.**
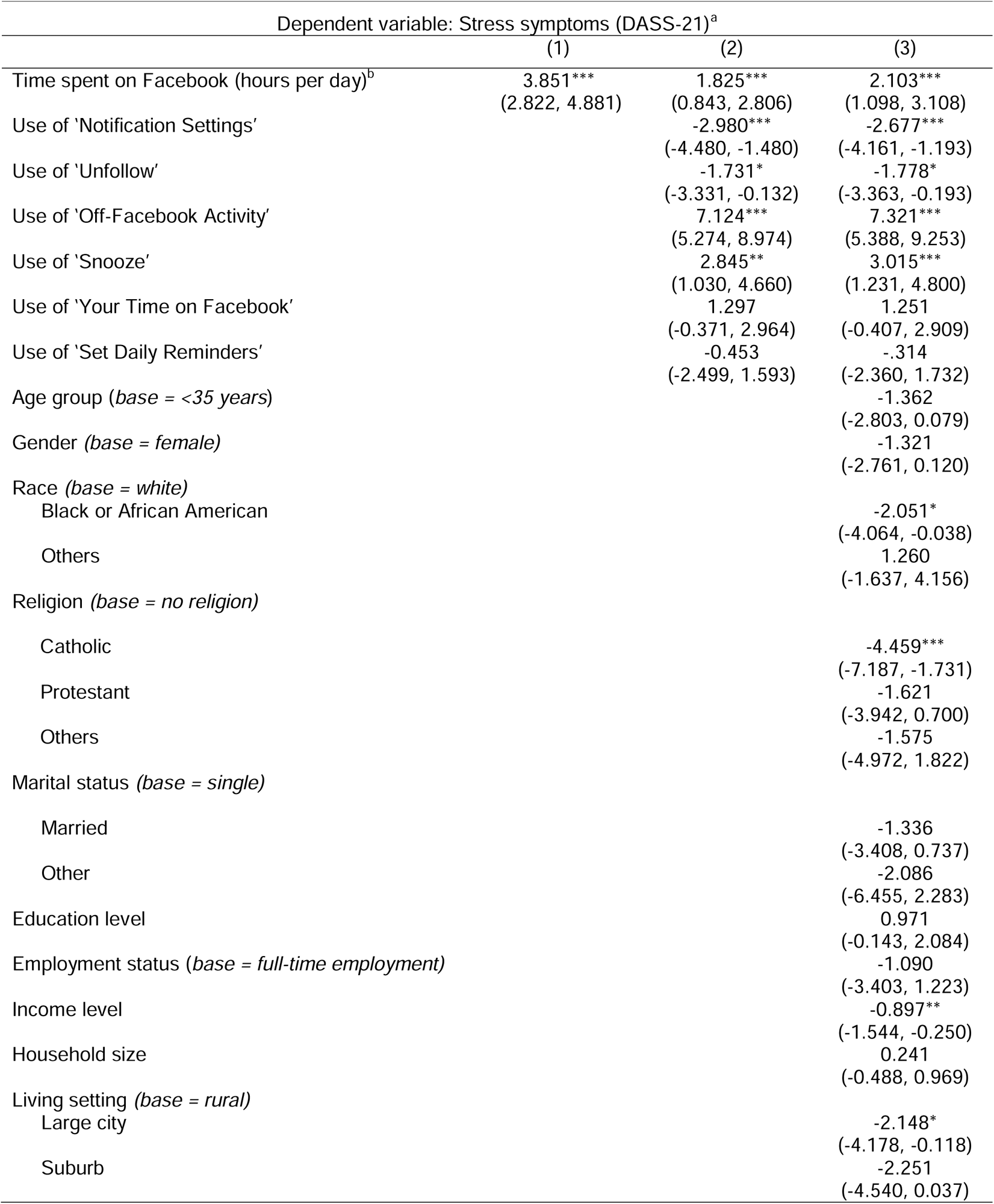
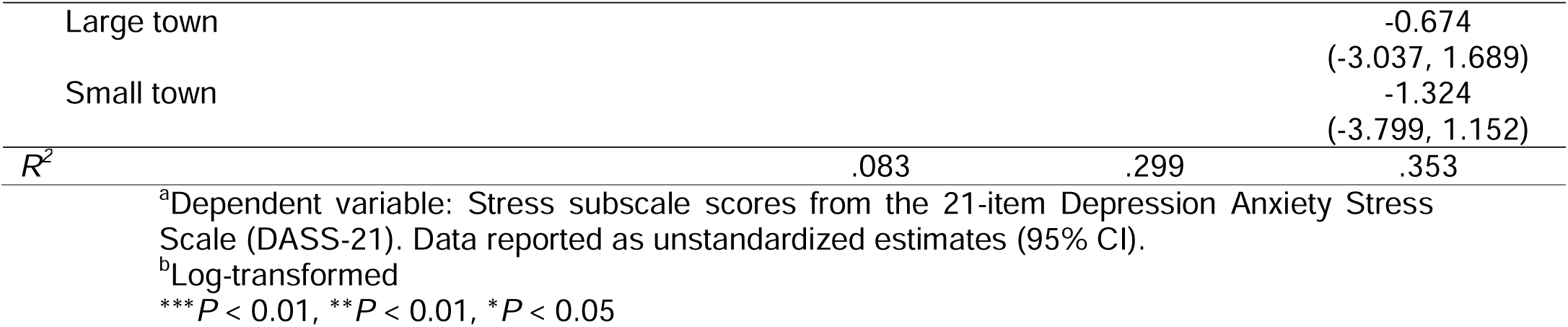
Predicting stress symptoms as a function of Facebook usage patterns.

When we added participants’ use of Facebook features as predictors, the amount of variance accounted from increased from 8.3% (Model 1) to 29.9% (Model 2; F(6, 587)=30.154, R^2^=0.216, *P*<0.001). Once again, we found that participants who used the Notification Shortcut Bar (β=2.980, *P*<0.001) and Unfollow button (β=-1.731, *P*=0.034) showed lower stress scores, but those who used Off-Facebook Activity (β=-7.124, *P*<0.001) and the Snooze button (β=2.845, *P*=0.002) showed higher stress scores. These associations remained significant in Model 3, when demographic variables were controlled for. Across both Models 2 and 3, there were no significant associations between stress scores and use of either Your Time on Facebook or Set Time Reminder (smallest *P*=0.13).

#### 3.3.4. Sensitivity analyses

As sensitivity analyses, we repeated Models 2 and 3 for each outcome variable without the inclusion of participants’ duration of Facebook use. As shown in Appendix 1 (Tables S1-S3), the key findings pertaining to Facebook’s digital well-being tools did not change.

## 4. Discussion

In this paper, we presented the first empirical study of Facebook’s digital well-being tools. First, echoing prior studies [8], we found that participants who spent more time on Facebook had more symptoms of depression, anxiety, and stress. Accounting for Facebook consumption alone explained a tenth of the variance (8-13%) in participants’ well-being. Consequently, we examined: (i) whether participants used the platform’s digital well-being tools, and (ii) whether usage was associated with better mental health.

Although most participants (97%) knew about Facebook’s well-being tools, each tool was used by only 17 to 55% of participants, and largely on an ad-hoc basis. In turn, participants who used either the Notification Settings feature or Unfollow tools reported fewer symptoms of depression, anxiety, and stress. Conversely, those who used either Snooze or Off-Facebook Activity showed higher scores on each of these measures. Finally, there was no evidence that Your Time on Facebook or Set Daily Reminders was associated with well-being. This set of findings was robust, observed regardless of whether we controlled for participants’ duration of Facebook use or their sociodemographic factors.

Taken together, our findings underscore the complexity of designing social media platforms to optimize user welfare. Out of the six digital well-being tools we examined, only two were associated with a decreased risk for mental health symptoms: (i) a feature toning down the amount of content brought to the user’s attention (Notification Shortcut Bar); and (ii) a feature allowing users to customize their newsfeeds, minimizing social comparison (Unfollow). Nonetheless, it remains unclear why two other features that supported customization of newsfeeds (Snooze and Off-Facebook Activity) predicted a *higher* risk for mental health symptoms. Further research is thus needed to understand these patterns.

It is noteworthy that we found no significant association between the use of time-monitoring features (Your Time on Facebook, Set Daily Reminders) and well-being. This finding is counterintuitive because time spent on Facebook has been linked repeatedly to poorer mental health outcomes (including in this study) [8]. Consequently, most social media developers have incorporated time-monitoring features into their digital well-being programs,

allowing users to track how much time they have spent on a platform or to set limits on usage (e.g., on YouTube, Instagram, Facebook, TikTok) [13, 16, 22]. Nonetheless, we found no empirical support for this widely-used strategy – consistent with ‘digital detox’ studies reporting that interventions to curb social media use have a limited impact on mood and well-being [23].

### 4.1 Implications

Moving forward, our study has several implications for research and practice. First, it appears that current well-being measures by social media platforms may be insufficient. This begs the question of how digital well-being tools should be designed to maximize user benefits. Despite widespread calls for app developers to prioritize their users, there remains limited empirical data to guide platforms in carrying out this mandate. We thus urge researchers to address this gap, allowing an evidence-based toolkit of in-app well-being features to be developed.

### 4.2 Limitations

In reporting our findings, we note several limitations of our study. First, we chose the design of an epidemiological survey [24, 25]. In a new area of research, this allowed us to: (i) document baseline adoption rates for digital well-being tools, and (ii) to examine multiple tools at the same time. Nonetheless, correlation does not equate to causation, and our findings need to be followed up with randomized controlled trials. Second, we recruited participants within the general population of internet users. It is possible that stronger effects would be observed in vulnerable groups – for example, amongst individuals with problematic forms of Facebook usage [26], or amongst adolescents. Further research should thus explore this possibility. Finally, we focused on Facebook because of its widespread popularity. It is currently unclear whether our findings would generalize to other social networking services (e.g., Instagram).

### 4.3 Conclusion

In the 2022 State of the Union Address, President Joe Biden called for social media platforms to be held accountable, for companies to pursue user benefits over profits [27]. Amid these petitions, there is a need to understand how social media platforms can be designed to optimize user well-being. Accordingly, our study provides the first line of evidence that two digital well-being features may be linked to improved mental health. At the same time, we also caution app developers that: (i) not all well-being features are alike, and indeed that (ii) certain features could backfire. Moving forward, we urge further research to develop and carefully investigate the impact of digital well-being tools on social media.

## Data Availability

All data produced in the present study are available upon reasonable request to the authors.

## Acknowledgements

This research was funded by a grant awarded to JCJL from the National University of Singapore’s Centre for Trusted Internet and Community (CTIC-RP-20-09). The third author’s involvement (ARM) was funded by a centre grant awarded to the Centre for Sleep and Cognition.

