## Appendix1 for "Predicting psychological symptoms when Facebook’s digital well-being features are used: A cross-sectional survey"

**Appendix 1**

In Tables S1-S3, we ran two additional models to predict the DASS-21 subscale scores (depression, anxiety, and stress). In Model A, we included the 6 variables pertaining to participants’ use of each Facebook digital well-being tool. In Model B, Model A was repeated with the inclusion of demographic variables. Our conclusions pertaining to the Facebook well-being tools did not change.

*Table S1.* Predicting depression symptoms as a function of Facebook usage patterns.

| Dependent variable: Depression symptoms (DASS-21)^a^ | | |
| --- | --- | --- |
|  | (A)^b^ | (B)^b^ |
| Use of ‘Notification Settings’ | -1.561**  (-2.622,-.0499) | -1.369*  (-2.429,-0.310) |
| Use of ‘Unfollow’ | -1.075  (-2.206, 0.056) | -0.983  (-2.112, 0.147) |
| Use of ‘Off-Facebook Activity’ | 5.429***  (4.151, 6.707) | 5.588***  (4.218, 6.958) |
| Use of ‘Snooze’ | 2.442***  (1.153, 3.731) | 2.479***  (1.201, 3.757) |
| Use of ‘Your Time on Facebook’ | 0.638  (-0.542,1.819) | 0.550  (-0.635, 1.736) |
| Use of ‘Set Daily Reminders’ | 0.155  (-1.291, 1.601) | 0.079  (-1.377, 1.535) |
| Age group (*base = <35 years*) |  | -0.758  (-1.789, 0.273) |
| Gender *(base = female)* |  | -0.618  (-1.649, 0.413) |
| Race *(base = white)* |  |  |
| Black or African American |  | -1.406  (-2.840, 0.029) |
| Others |  | -0.408  (-2.475, 1.660) |
| Religion *(base = no religion)* |  |  |
| Catholic |  | -2.518**  (-4.456, -0.580) |
| Protestant |  | -0.828  (-2.472, 0.816) |
| Others |  | -1.027  (-3.404, 1.350) |
| Marital status *(base = single*) |  |  |
| Married |  | -0.594  (-2.064, 0.876) |
| Others |  | 1.149  (-1.978, 4.276) |
| Education level |  | 0.526  (-0.269, 1.321) |
| Employment status *(base = full-time employment)* |  | -0.587  (-2.232, 1.059) |
| Income level |  | -0.745**  (-1.209, -0.281) |
| Household size |  | 0.335  (-0.182, 0.851) |
| Living setting *(base = rural)* |  |  |
| Large city |  | -1.687*  (-3.134, -0.240) |
| Suburb |  | -1.608  (-3.239, 0.022) |
| Large town |  | -0.848  (-2.536, 0.840) |
| Small town |  | -1.589  (-3.348, 0.169) |
| *R^2^* | 0.274*** | 0.319*** |

^a^Dependent variable: depression subscale scores from the 21-item Depression Anxiety Stress Scale (DASS-21). Data reported as beta estimates (95% CI).

^b^Models A and B correspond to Models 2 and 3 of Table 3, respectively.

****P* < 0.01, ***P* < 0.01, **P* < 0.05

*Table S2.* Predicting anxiety symptoms as a function of Facebook usage patterns.

| Dependent variable: Anxiety symptoms (DASS-21)^a^ | | |
| --- | --- | --- |
|  | (A)^b^ | (B)^b^ |
| Use of ‘Notification Settings’ | -2.337***  (-3.572, -1.101) | -1.993***  (-3.213, -0.773) |
| Use of ‘Unfollow’ | -1.213  (-2.530, 0.103) | -1.231  (-2.532, 0.069) |
| Use of ‘Off-Facebook Activity’ | 7.627***  (6.139, 9.114) | 6.596***  (5.019, 8.173) |
| Use of ‘Snooze’ | 3.305***  (1.805, 4.805) | 3.434***  (1.963, 4.905) |
| Use of ‘Your Time on Facebook’ | 1.574*  (0.200, 2.948) | 1.097  (-0.267, 2.462) |
| Use of ‘Set Daily Reminders’ | 0.414  (-1.268, 2.097) | 0.016  (-1.661, 1.692) |
| Age group (*base = <35 years*) |  | -1.031  (-2.218, 0.157) |
| Gender *(base = female)* |  | -1.124  (-2.311, 0.063) |
| Race *(base = white)* |  |  |
| Black or African American |  | -0.821  (-2.473, 0.830) |
| Others |  | -1.093  (-3.474, 1.287) |
| Religion *(base = no religion)* |  |  |
| Catholic |  | -1.045  (-3.276, 1.187) |
| Protestant |  | 0.794  (-1.099, 2.687) |
| Others |  | 0.521  (-2.216, 3.259) |
| Marital *(base = single)* |  |  |
| Married |  | -0.080  (-1.773, 1.612) |
| Others |  | -3.433  (-7.033, 0.167) |
| Education level |  | 0.913  (-0.003, 1.828) |
| Employment status *(base = full-time employment)* |  | -1.465  (-3.360, 0.430) |
| Income level |  | -0.733**  (-1.267, -0.199) |
| Household size |  | 0.465  (-0.130, 1.059) |
| Living setting *(base = rural)* |  |  |
| Large city |  | -2.213**  (-3.879, -0.547) |
| Suburb |  | -2.829**  (-4.707, -0.952) |
| Large town |  | -1.216  (-3.159, 0.727) |
| Small town |  | -1.741  (-3.765, 0.283) |
| *R^2^* | 0.371*** | 0.423*** |

^a^Dependent variable: Anxiety subscale scores from the 21-item Depression Anxiety Stress Scale (DASS-21). Data reported as beta estimates (95% CI).

^b^Models A and B correspond to Models 2 and 3 of Table 3, respectively.

****P* < 0.01, ***P* < 0.01, **P* < 0.05

*Table S3.* Predicting stress symptoms as a function of Facebook usage patterns.

| Dependent variable: Stress symptoms (DASS-21)^a^ | | |
| --- | --- | --- |
|  | (A)^b^ | (B)^b^ |
| Use of ‘Notification Settings’ | -2.989***  (-4.499, -1.479) | -2.636***  (-4.138, -1.135) |
| Use of ‘Unfollow’ | -1.411  (-3.019, 0.197) | -1.431  (-3.032, 0.170) |
| Use of ‘Off-Facebook Activity’ | 7.747***  (5.929, 9.564) | 7.688***  (5.747, 9.630) |
| Use of ‘Snooze’ | 3.016***  (1.183, 4.849) | 3.137***  (1.326, 4.948) |
| Use of ‘Your Time on Facebook’ | 1.604  (-0.074, 3.283) | 1.469  (-0.211, 3.148) |
| Use of ‘Set Daily Reminders’ | -0.298  (-2.354, 1.758) | -0.302  (-2.366, 1.762) |
| Age group (*base = <35 years*) |  | -1.377  (-2.839, 0.084) |
| Gender *(base = female)* |  | -1.259  (-2.720, 0.202) |
| Race *(base = white)* |  |  |
| Black or African American |  | -1.662  (-3.695, 0.371) |
| Others |  | 0.667  (-2.263, 3.596) |
| Religion *(base = no religion)* |  |  |
| Catholic |  | -3.669**  (-6.415, -0.922) |
| Protestant |  | -0.892  (-3.221, 1.438) |
| Others |  | -0.019  (-3.388, 3.350) |
| Marital status *(base = single)* |  |  |
| Married |  | -0.692  (-2.775, 1.391) |
| Others |  | -1.677  (-6.109, 2.754) |
| Education level |  | 1.024  (-0.103, 2.151) |
| Employment status *(base = full-time employment)* |  | -0.953  (-3.285, 1.380) |
| Income level |  | -0.891**  (-1.549, -0.234) |
| Household size |  | 0.282  (-0.450, 1.014) |
| Living setting *(base = rural)* |  |  |
| Large city |  | -2.442*  (-4.493, -0.391) |
| Suburb |  | -2.717*  (-5.028, -0.407) |
| Large town |  | -0.916  (-3.308, 1.475) |
| Small town |  | -1.740  (-4.232, 0.752) |
| *R^2^* | 0.283** | 0.333*** |

^a^Dependent variable: Stress subscale scores from the 21-item Depression Anxiety Stress Scale (DASS-21). Data reported as beta estimates (95% CI).

^b^Models A and B correspond to Models 2 and 3 of Table 3, respectively.

****P* < 0.01, ***P* < 0.01, **P* < 0.05
